# Reliability and Validity of the Dutch Interoceptive Accuracy Scale and Interoceptive Attention Scale

**DOI:** 10.1101/2025.05.06.25326009

**Authors:** Jesper Mulder, Marije T. Elferink-Gemser, Juriena D. de Vries, Jessica C. Kiefte-de Jong

## Abstract

Interoception, the perception of internal bodily signals, is thought to play a key role in health behavior. Interoceptive domains (e.g., accuracy and attention) are associated with mental disorders in various ways, underscoring the need for specific measures for different interoceptive domains. To allow for studying interoceptive domains in the Dutch population, the current study aimed to investigate the reliability and validity of Dutch translations of the Interoceptive Accuracy Scale (IAS-D) and Interoceptive Attention Scale (IATS-D).

In a sample of 779 participants (mean age 52.98 ± 16.47, 49.6% female), the IAS-D showed good internal consistency (α = 0.89) with a four-factor structure according to the confirmatory factor analysis (CFI = 0.816, TLI = 0.788, RMSEA = 0.084). The IAS-D had significant negative relations to the Interoceptive Confusion Questionnaire (β = −0.638, p < 0.001), depression (β = −0.298, p < 0.001), and alexithymia (β = 0.528, p < 0.001). The IATS-D showed good internal consistency (α = 0.94) with a three-factor structure according to the confirmatory factor analysis (CFI = 0.852, TLI = 0.833, RMSEA = 0.100). The IATS-D had significant positive relations with the Interoceptive Confusion Questionnaire (β = 0.481, p < 0.001), Body Perception Questionnaire (β = 0.207, p < 0.001), depression (β = 0.377, p < 0.001), and alexithymia (β = 0.528, p < 0.001).

The IAS-D and IATS-D are considered reliable and valid instruments for assessing self-reported interoceptive accuracy and attention in Dutch-speaking populations, supporting public health research on the role of interoception in health behavior.

**Highlights:**

- Interoceptive domains associate with mental disorders in various ways
- Specific measures for interoceptive domains are needed
- IAS-D and IATS-D are reliable and valid instruments
- IAS-D and IATS-D can distinguish between interoceptive domains

## 1. Introduction

Interoception is defined as the perception of the internal state of the body, including a wide range of physical states like heart rate, respiration, temperature, fatigue, hunger, satiety, muscle ache, and pain (Craig, 2002, 2003, 2009; Murphy et al., 2019). Research in the field of interoception has increased over the years, which has provided a better understanding of the important role of interoception in physical and mental well-being. Interoception helps in maintaining homeostasis (i.e., the process of maintaining a stable internal environment) and allostasis (i.e., the process of actively adjusting and adapting to changing conditions). Changes in internal bodily states can also affect behaviors (Critchley & Harrison, 2013; Farb et al., 2015; Quadt et al., 2018). Several mental disorders have been associated with impaired interoception, such as depressive disorders and alexithymia (i.e., an inability to recognize or describe one’s own emotions). Lower scores on interoceptive measures have been related to more depressive symptoms (Pollatos et al., 2009; Quadt et al., 2018), and higher alexithymia scores (Brewer et al., 2016; Murphy et al., 2020). However, interoceptive domains (e.g., accuracy or attention) and modalities (e.g., heartrate or respiration) are associated with these clinical symptoms in different ways. For instance, in anxiety respiratory interoceptive accuracy is decreased, while thermoceptive interoceptive accuracy is increased (Schoeller et al., 2025). This shows the complexity of interoception and the need for specific measures for different interoceptive domains and modalities.

Definitions and measures of interoceptive domains and modalities vary greatly across studies (Forkmann et al., 2016). Therefore, it is necessary to clearly define these domains in order to accurately describe interoception, while also acknowledging its multimodal and multifaceted nature. A promising model in this regard is the 2×2 factorial structure for interoception (Figure 1) developed by Murphy and colleagues (Murphy et al., 2019). This model suggests two factors. The first factor makes the distinction between two domains: interoceptive accuracy (i.e., an individual’s ability to perceive interoceptive signals accurately) and attention (i.e., the degree to which interoceptive signals are objects of attention). The second factor distinguishes between the way these domains (i.e., interoceptive accuracy or attention) is measured: an objective performance measure or a self-report measure. Measuring different facets of interoception may provide more accurate assessments (Schoeller et al., 2025). In the current study we will focus on self-reported interoception, as this allows us to investigate interoception in large samples. Based on the 2×2 factorial model of interoception, two self-report measures of interoception have been developed in general populations: the Interoceptive Accuracy Scale (IAS; Murphy et al., 2020) and the Interoceptive Attention Scale (IATS; Gabriele et al., 2022). These two questionnaires were developed as unidimensional measures in general populations, while existing questionnaires such as the Interoceptive Confusion Questionnaire (ICQ) and Body Perception Questionnaire (BPQ) are often multidimensional and more tailored to clinical dysfunction or broad autonomic nervous system awareness. Since the accuracy and attention domains of interoception seem to be distinct but complementary (Brand et al., 2023; Gabriele et al., 2022; Murphy et al., 2020; Murphy et al., 2019; Tünte et al., 2024), and questionnaires focusing on these domains are not yet available in Dutch, there is interest to investigate the psychometric properties of Dutch versions of the IAS and IATS.

**Figure 1.**
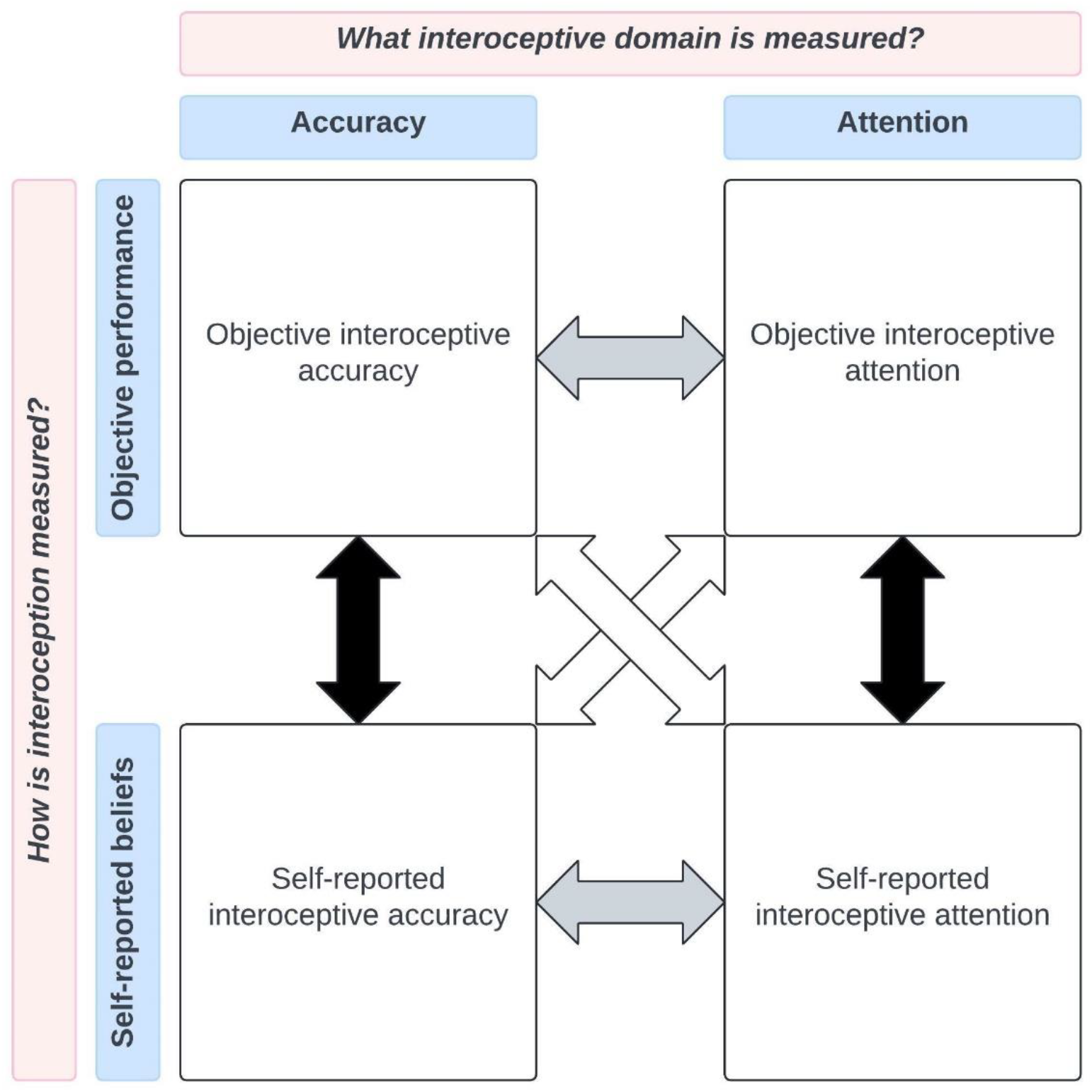
Simplified version of the 2×2 factorial structure for interoception developed by Murphy et al. (Murphy et al., 2019). The quadrants represent interoceptive accuracy assessed with objective performance tasks (top left), interoceptive attention measured with objective performance tasks (top right), interoceptive accuracy assessed with self-report measures (bottom left), and interoceptive attention assessed with self-report measures (bottom right). The arrows represent the interaction between objective and self-report measures (i.e., interoceptive awareness; black arrows), interoceptive accuracy and attention (grey arrows), and across different interoceptive domains and measures (white arrows).

The objective of the current study is to investigate the psychometric properties of Dutch translations of the IAS and IATS (IAS-D and IATS-D) in a sample of the general Dutch population. We will investigate the internal consistency, factor structures, and convergent, discriminant, and criterion validity of the questionnaires. Consequently, we compare the IAS-D and IATS-D to validated questionnaires: the ICQ, BPQ Short Form (BPQ-SF), and questionnaires on depression and alexithymia. We will base our hypotheses on the findings of previous studies (Brand et al., 2023; Gabriele et al., 2022; Murphy et al., 2020; Murphy et al., 2019; Tünte et al., 2024): the IAS is expected to have a significant negative relation with the ICQ, and the IATS is expected to have a significant positive relation with both the ICQ and BPQ-SF. We expect no significant relation between the IAS and BPQ-SF. To investigate criterion validity the correlation between the IAS, IATS, depression, and alexithymia will be examined. We hypothesize that interoceptive accuracy relates negatively, and interoceptive attention positively, to depressive symptoms (Brand et al., 2023; Tünte et al., 2024). It is also expected that interoceptive accuracy negatively relates to alexithymia, and interoceptive attention positively to alexithymia (Murphy et al., 2020; Tünte et al., 2024).

## 2. Methods

### 2.1. Population

This study targeted a population that gives a fair representation of the general Dutch population, based on sex, age, region/province, and socio-economic status. To be eligible to participate in the study, a subject had to meet the following criteria: the subject was 1) at least 18 years old, and 2) had a clear understanding of the Dutch language. Participants in this study were recruited through a Dutch internet panel, administered by research agency Flycatcher (www.flycatcher.eu). This panel consists of 20.000 members from the Dutch public, thus providing a fair representation of the general Dutch population. Participants voluntarily and actively indicated their willingness to participate in the online surveys, through Flycatcher’s ‘double-active-opt-in’. Flycatcher meets high quality requirements and is ISO-certified. Sample size calculation using G*Power 3.1 (Faul et al., 2009) indicated a minimum sample size of 812 participants. The participants of this study were contacted via email to take part in this study. Participants were given points that could be spend on (charity) gift vouchers for completing the surveys.

### 2.2. Measures

#### 2.2.1. Self-reported interoceptive accuracy

The Interoceptive Accuracy Scale (IAS; Murphy et al., 2020) is a questionnaire constructed to assess individuals’ accuracy in evaluating sensations that have either been described as interoceptive or are associated with activation in the insula, an area commonly associated with the processing of interoceptive signals. An example of a question in this questionnaire is “I can always accurately perceive when my heart is beating fast.” The scale comprises of 21 items rated on a scale from strongly disagree (1) to strongly agree (5), with total scores ranging from 21 to 105. Higher scores indicate greater self-reported interoceptive accuracy. English and German versions of the IAS (α = 0.84 to 0.90, ω = 0.81 to 0.88; Brand et al., 2023; Gabriele et al., 2022; Murphy et al., 2020; Tünte et al., 2024) have been shown to have good to excellent internal consistency.

The Interoceptive Confusion Questionnaire (ICQ; Brewer et al., 2016) assesses the degree to which individuals feel that they struggle to interpret their own non-affective interoceptive states, such as hunger, temperature and arousal. Examples of questions in this questionnaire are “I am very sensitive to changes in my heartrate” and “When I adjust the heat of a room or car, others find it uncomfortable.” Items are scored on a scale from 1 (“Does not describe me”) to 5 (“Describes me very well”). Total scores range between 20 and 100. Higher scores indicate greater self-reported interoceptive confusion. The ICQ has low to acceptable internal consistency (ω = 0.66 to 0.75, α = 0.44 to 0.53; Brand et al., 2023; Brewer et al., 2016; Gabriele et al., 2022; Murphy et al., 2020; Tünte et al., 2024), which is considered adequate for this study.

#### 2.2.2. Self-reported interoceptive attention

The Interoceptive Attention Scale (IATS; Gabriele et al., 2022) is a questionnaire constructed to quantify the extent to which internal signals are the object of one’s attention. The signals exactly match the signals included in the IAS. An example of a question in this questionnaire is “Most of the time my attention is focused on whether my heart is beating fast.” The IATS comprises 21 items rated from strongly disagree (1) to strongly agree (5), with total scores ranging from 21 to 105. Higher scores indicate greater self-reported attention to internal signals. The IATS (α = 0.91, ω = 0.85 to 0.92; Gabriele et al., 2022; Tünte et al., 2024) has been shown to have good to excellent internal consistency in English and German speaking populations.

The Body Perception Questionnaire Short Form (BPQ-SF; Cabrera et al., 2018) was developed to assess the subjective experiences of the function and reactivity of target organs and structures that are innervated by the autonomic nervous system. The original BPQ has 122 items and assesses body awareness, autonomic nervous system reactivity, cognitive-emotional-somatic stress response, body and cognitive stress response styles, and health history. The short form focuses on the awareness and autonomic reactivity subscales of the BPQ, resulting in 46 items scored from never (1) to always (5). Total scores range between 46 and 230. Example items of the BPQ-SF are “During most situations I am aware of swallowing frequently” and “During most situations I am aware of how hard my heart is beating.” Higher scores indicate greater self-reported interoceptive attention. The BPQ-SF has good to excellent internal consistency across various subscales (ω = 0.77 to 0.98; Brand et al., 2023; Cabrera et al., 2018; Gabriele et al., 2022; Murphy et al., 2020; Tünte et al., 2024). In German samples the BPQ-SF was found to be significantly related to the IAS (Brand et al., 2023; Tünte et al., 2024), but in English samples this was not the case (Gabriele et al., 2022; Murphy et al., 2020). The BPQ-SF was found to be significantly related to the IATS (Gabriele et al., 2022; Tünte et al., 2024).

#### 2.2.3. Depression

The Beck Depression Inventory (BDI-II; Beck et al., 1996) measures the severity of depressive symptoms. The questionnaire consists of 21 groups of statements assessing the presence of psychological and physiological symptoms of major depression. Statements are assigned point values (ranging from 0 to 3) reflecting the severity of depressive symptoms. Total scores range from 0 to 63, with a higher score indicating more depressive symptoms. Example answers from an item are: “I do not feel sad”, “I feel sad”, “I am sad all the time and I cannot snap out of it”, or “I am so sad and unhappy that I cannot stand it.” The internal consistency of the Dutch BDI-II has been reported as Cronbach’s α = 0.95 (Roelofs et al., 2013).

#### 2.2.4. Alexithymia

The Toronto Alexithymia Scale (TAS-20; Bagby et al., 1994) is a 20-item scale to assess alexithymia traits. The scale includes items like “I am often confused about what emotion I am feeling” and “I have physical sensations that even doctors do not understand.” The scale is rated on a 5-point forced-choice answer format ranging from strongly disagree (1) to strongly agree (5), and grouped in three subscales: difficulty identifying feelings, difficulty describing feelings, and externally oriented thinking. Higher scores indicate stronger alexithymia traits. Total scores range from 20 to 100. The internal consistency of Dutch versions of the TAS-20 range from Cronbach’s α = 0.79 to 0.83 (Kooiman et al., 2002; Veehof et al., 2011).

### 2.3. Procedures

The IAS and IATS questionnaires were translated to Dutch, using cross-cultural guidelines (Sousa & Rojjanasrirat, 2011), resulting in the IAS-D and IATS-D. The translation was performed by an organization specializing in medical translations. After translation, the Dutch questionnaires were evaluated in individual think-aloud interviews with a small group of Dutch speaking subjects from the general population. Where necessary, adjustments were made to the Dutch versions of the questionnaires based on feedback from the think-aloud interviews (Appendix A). Adjustments were discussed with the other researchers until consensus was reached, resulting in a final version of both questionnaires. The ICQ was not available in Dutch and was translated using the same method as described above, however this questionnaire was not validated in the current study.

### 2.4. Statistical analyses and software

All analyses were performed in Python version 3.12, using the factor-analyzer (version 0.5), scikit-learn (version 1.5), scipy (version 1.14), semopy (version 2.3), and statsmodels (version 0.14) packages. Normality of data for all measures was checked beforehand, by visually examining the distribution of data using histograms and Q-Q plots, and performing the Shapiro-Wilk test. Descriptive statistics (mean, standard deviation, median, interquartile range, minimum, and maximum) were calculated to summarize the characteristics and questionnaire scores of the participants. Cronbach’s alpha was calculated, an alpha of 0.7 or higher was considered acceptable (Gliem & Gliem, 2003). Initially, correlations between items were examined, the Kaiser-Meyer-Olkin (KMO) measure of sampling adequacy and Bartlett’s test of sphericity were calculated, and communalities were checked. For the KMO, values above 0.5 are recommended (Williams et al., 2010). Next, exploratory principal components analyses (PCA) and confirmatory factor analyses (CFA) were performed to identify underlying structures of the Dutch IAS and IATS, and compare these structures to factor structures identified in previous studies. The PCA was performed with either orthogonal or oblique rotation, depending on the correlation between factors. The PCA was performed on all available IAS and IATS data. To confirm the structure of the Dutch IAS and IATS as identified in the PCA, a confirmatory factor analysis (CFA) was performed. The fit indices comparative fit index (CFI), Tucker-Lewis index (TLI), and root mean square error of approximation (RMSEA) were calculated. For the CFI and TLI values above 0.90 are usually considered to be indicators of good fit (Hooper et al., 2007). For the RMSEA values between 0.06 and 0.10 indicate acceptable fit (Hooper et al., 2007).

Convergent, discriminant, and criterion validity were determined using linear regression analyses. The dependent variables were the total scores of the IAS and IATS. The independent variables were the total scores of the ICQ, BPQ-SF, TAS-20, and BDI-II. For convergent validity the IAS was compared to the ICQ, and the IATS to the BPQ-SF. For discriminant validity the IAS was compared to the BPQ-SF, and the IATS to the ICQ. To investigate criterion validity the directions of the relations between the IAS, IATS, TAS-20, and BDI-II were investigated. In all analyses, alpha was set at 0.05 a priori.

## 3. Results

In total, 815 participants responded to the questionnaires. After removal of outliers (i.e., ≥ three standard deviations), 779 participants were included in the analyses. The remaining participants consisted of 393 (50.4%) males and 386 (49.6%) females, with an average age of 52.98 (SD = 16.47) years. Our sample showed similar scores on interoceptive measures, but results indicated more depressive symptoms and stronger alexithymia traits, compared to previous studies using the same questionnaires. Table 1 shows the average score per questionnaire.

**Table 1.** Mean age and average total scores per questionnaire based on 779 participants.

| Variable | Mean | SD | Median | Q1 | Q3 | Min | Max |
| --- | --- | --- | --- | --- | --- | --- | --- |
| Age | 52.98 | 16.47 | 54 | 41 | 67 | 19 | 92 |
| *Interoceptive accuracy* |  |  |  |  |  |  |  |
| IAS-D | 80.01 | 9.39 | 80 | 75 | 84 | 50 | 105 |
| ICQ | 47.36 | 7.18 | 47 | 42 | 53 | 27 | 66 |
| *Interoceptive attention* |  |  |  |  |  |  |  |
| IATS-D | 50.58 | 14.72 | 49 | 41 | 62 | 21 | 86 |
| BPQ-SF | 101.02 | 25.02 | 100 | 82 | 120 | 46 | 172 |
| *Depression and alexithymia* |  |  |  |  |  |  |  |
| BDI-II | 28.42 | 6.37 | 27 | 24 | 32 | 21 | 52 |
| TAS-20 | 55.08 | 7.83 | 55 | 50 | 60 | 33 | 79 |

Note. IAS-D = Interoceptive Accuracy Scale Dutch version; ICQ = Interoceptive Confusion Questionnaire; IATS-D = Interoceptive Attention Scale Dutch version; BPQ-SF = Body Perception Questionnaire Short Form; BDI-II = Beck Depression Inventory; TAS-20 = Toronto Alexithymia Scale.

### 3.1. Interoceptive Accuracy Scale factor structure

Cronbach’s alpha indicated good internal consistency for the Dutch IAS (α = 0.89). All 21 items of the IAS-D showed weak to strong (0.2 < |r| < 0.79) correlations with another item. The KMO measure of sampling adequacy was 0.90, which was above the recommend value of 0.5 (Williams et al., 2010). Bartlett’s test of sphericity was significant (χ^2^ = 5639.77, p < 0.001), indicating significant correlations among variables.

A PCA with varimax rotation was used to examine the factor structure of the IAS-D. The scree plot identified a one-factor structure, but it should be noted that four factors with an eigenvalue bigger than 1 were found. Cumulative variance of these four factors showed that 33% of the variance was explained by the first factor, while the following three factors together accounted for 19%. This prominence of the first factor was also seen in its eigenvalue of 6.96, compared to the next biggest eigenvalue of 1.55. All communalities were close to 1, indicating all items were strongly associated with the underlying factors. A CFA was performed to evaluate the fit of the model. Based on the findings of the PCA, goodness of fit indices for several models were tested (Table 2). These analyses indicated a four-factor model to be the best fit. Table 3 shows the factor loadings of the IAS-D items based on this four-factor model.

**Table 2.** CFA goodness of fit indices for the one- to four-factor models for the Interoceptive Accuracy Scale.

| **Model** | **CFI** | **TLI** | **RMSEA** |
| --- | --- | --- | --- |
| One-factor model | 0.756 | 0.729 | 0.096 |
| Two-factor model | 0.780 | 0.754 | 0.091 |
| Three-factor model | 0.792 | 0.765 | 0.089 |
| Four-factor model | 0.816 | 0.788 | 0.084 |

**Table 3.** Factor loadings for the Interoceptive Accuracy Scale items, based on the four-factor structure identified in the CFA.

| **Item** | **Factor 1** | **Factor 2** | **Factor 3** | **Factor 4** |
| --- | --- | --- | --- | --- |
| Item 7: Tasting | 1 |  |  |  |
| Item 8: Vomiting | 1.408 |  |  |  |
| Item 9: Sneezing | 1.564 |  |  |  |
| Item 10: Coughing | 1.470 |  |  |  |
| Item 11: Temperature | 1.217 |  |  |  |
| Item 12: Sexual arousal | 1.298 |  |  |  |
| Item 15: Muscle soreness | 1.375 |  |  |  |
| Item 17: Pain | 1.356 |  |  |  |
| Item 1: Heart |  | 1 |  |  |
| Item 3: Breathing |  | 0.898 |  |  |
| Item 16: Bruising |  | 1.111 |  |  |
| Item 18: Blood sugar |  | 1.077 |  |  |
| Item 13: Wind |  |  | 1 |  |
| Item 14: Burping |  |  | 1.011 |  |
| Item 19: Affective touch |  |  | 0.758 |  |
| Item 20: Tickling |  |  | 0.698 |  |
| Item 21: Itching |  |  | 0.823 |  |
| Item 2: Hunger |  |  |  | 1 |
| Item 4: Thirst |  |  |  | 0.923 |
| Item 5: Urination |  |  |  | 0.932 |
| Item 6: Defecation |  |  |  | 0.816 |

### 3.2. Interoceptive Attention Scale factor structure

Cronbach’s alpha indicated good internal consistency for the Dutch IATS (α = 0.94). All 21 items of the IATS-D showed weak to strong (0.2 < |r| < 0.79) correlations with another item. The KMO measure of sampling adequacy was 0.95, which was above the recommend value of 0.5 (Williams et al., 2010). Bartlett’s test of sphericity was significant (χ^2^ = 9516.87, p < 0.001), indicating significant relations among variables.

A PCA with varimax rotation was performed to examine the factor structure of the IATS-D. As with the accuracy scale, for the IATS-D a one-factor structure was identified. This was indicated by the scree plot, the eigenvalue of the first factor (9.83), and the explained variance of the first factor (47%). All communalities were close to 1, indicating all items were strongly associated with the underlying factors. Based on the CFA fit indices evaluated for the one-, two-, three-, and four-factor models, it seemed the three-factor model is the best fit for the IATS-D (Table 4). Based on these findings, the factor loadings for the three-factor model are shown in Table 5.

**Table 4.** CFA goodness of fit indices for the one- to four factor models for the Interoceptive Attention Scale.

| **Model** | **CFI** | **TLI** | **RMSEA** |
| --- | --- | --- | --- |
| One-factor model | 0.835 | 0.817 | 0.105 |
| Two-factor model | 0.835 | 0.816 | 0.105 |
| Three-factor model | 0.852 | 0.833 | 0.100 |
| Four-factor model | 0.852 | 0.831 | 0.101 |

**Table 5.** Factor loadings for the Interoceptive Attention Scale items, based on the three-factor structure identified in the CFA.

| **Item** | **Factor 1** | **Factor 2** | **Factor 3** |
| --- | --- | --- | --- |
| Item 1: Heart | 1 |  |  |
| Item 3: Breathing | 1.082 |  |  |
| Item 8: Vomiting | 1.273 |  |  |
| Item 9: Sneezing | 1.451 |  |  |
| Item 10: Coughing | 1.391 |  |  |
| Item 11: Temperature | 1.192 |  |  |
| Item 12: Sexual arousal | 0.947 |  |  |
| Item 14: Burping | 1.309 |  |  |
| Item 15: Muscle soreness | 1.152 |  |  |
| Item 17: Pain | 1.233 |  |  |
| Item 18: Blood sugar | 1.044 |  |  |
| Item 21: Itching | 1.205 |  |  |
| Item 7: Tasting |  | 1 |  |
| Item 13: Wind |  | 3.396 |  |
| Item 19: Affective touch |  | 1.902 |  |
| Item 20: Tickling |  | 2.842 |  |
| Item 2: Hunger |  |  | 1 |
| Item 4: Thirst |  |  | 1.200 |
| Item 5: Urination |  |  | 1.421 |
| Item 6: Defecation |  |  | 1.608 |
| Item 16: Bruising |  |  | 1.188 |

### 3.3. Regression analyses

Univariate linear regression analyses were performed with the IAS-D as dependent variable, and the ICQ, BPQ-SF, BDI-II, and TAS-20 as independent variables. Our results indicated significant negative associations between the IAS-D scores and the ICQ (β = −0.638, 95% CI [−0.719, −0.558]), BDI-II (β = − 0.298, 95% CI [−0.399, −0.196]), and TAS-20 (β = −0.298, 95% CI [−0.380, −0.216]). Lower levels of interoceptive confusion, depressive symptoms, and alexithymia traits were associated with greater interoceptive accuracy. We observed no significant association between the IAS-D and the BPQ-SF (Table 6).

**Table 6.** Univariate regression analyses with the Interoceptive Accuracy Scale and Interoceptive Attention Scale as dependent variables, and the ICQ, BPQ-SF, BDI-II, and TAS-20 as independent variables.

| Variables | $\beta$ | SE | t | p | 95% CI |
| --- | --- | --- | --- | --- | --- |
| *Dependent variable: IAS-D* |  |  |  |  |  |
| ICQ | -0.638 | 0.041 | -15.568 | <0.001 | -0.719, -0.558 |
| BPQ-SF | 0.0002 | 0.013 | 0.017 | 0.986 | -0.026, 0.027 |
| BDI-II | -0.298 | 0.052 | -5.749 | <0.001 | -0.399, -0.196 |
| TAS-20 | -0.298 | 0.042 | -7.144 | <0.001 | -0.380, -0.216 |
| *Dependent variable: IATS-D* |  |  |  |  |  |
| ICQ | 0.481 | 0.072 | 6.725 | <0.001 | 0.341, 0.621 |
| BPQ-SF | 0.207 | 0.020 | 10.471 | <0.001 | 0.168, 0.246 |
| BDI-II | 0.377 | 0.082 | 4.606 | <0.001 | 0.216, 0.537 |
| TAS-20 | 0.527 | 0.065 | 8.147 | <0.001 | 0.400, 0.655 |

*Note.* IAS-D = Interoceptive Accuracy Scale Dutch version; ICQ = Interoceptive Confusion Questionnaire; IATS-D = Interoceptive Attention Scale Dutch version; BPQ-SF = Body Perception Questionnaire Short Form; BDI-II = Beck Depression Inventory; TAS-20 = Toronto Alexithymia Scale.

The same analyses were performed with the IATS-D as dependent variable. The ICQ (β = 0.481, 95% CI [0.341, 0.621]), BPQ-SF (β = 0.207, 95% CI [0.168, 0.246]), BDI-II (β = 0.377, 95% CI [0.216, 0.537]), and TAS-20 (β = 0.528, 95% CI [0.400, 0.655]) were significant determinants of IATS-D scores (Table 6). Higher levels of interoceptive confusion, autonomic nervous system reactivity, depressive symptoms, and alexithymia traits were associated with greater interoceptive attention.

## 4. Discussion

This study aimed to develop Dutch versions of the IAS (i.e., interoceptive accuracy; Murphy et al., 2020) and IATS (i.e., interoceptive attention; Gabriele et al., 2022), and to examine their internal consistency, factor structure, and convergent, discriminant, and criterion validity. The IAS-D showed good internal consistency. A four-factor structure was the best fit according to the CFA. The IATS-D also showed good internal consistency. For the IATS-D, a three-factor structure showed the best fit in the CFA. The regression analyses confirmed our hypotheses for both the IAS-D and IATS-D.

The first factor of the IAS-D (tasting, vomiting, sneezing, coughing, temperature, sexual arousal, muscle soreness, and pain) may capture acute bodily sensations related to discomfort, arousal, and physiological responses. The second factor (heart, breathing, bruising, and blood sugar) seems to relate to cardiovascular and respiratory internal physiological processes. The third factor (wind, burping, affective touch, tickling, and itching) may encompass tactile and gastrointestinal bodily sensations. Finally, the fourth factor (hunger, thirst, urination, and defecation) seem to consist of basic biological needs. In the two-factor solution found in the original IAS, the first factor was described as the perception of interoceptive signals (e.g., heartrate) and the second factor as signals difficult to perceive with interoceptive information alone (e.g., bruising indicated by skin discoloration; Murphy et al., 2020). In comparison to this two-factor structure, the current four-factor structure seems to spread the items out over more specific interoceptive modalities and signals.

Similarly, the IATS-D consisted of three factors, the first (heart, breathing, vomiting, sneezing, coughing, temperature, sexual arousal, burping, muscle soreness, pain, blood sugar, and pain) of which may encompass internal bodily sensations related to physiological discomfort, autonomic processes, and arousal states. The second factor (tasting, wind, affective touch, and tickling) consists of tactile and gastrointestinal bodily sensations. The final factor (hunger, thirst, urination, defecation, and bruising) seems to indicate basic physiological needs. Based on the CFA, the IAS-D and IATS-D seem to show similar underlying structures. However, in the three-factor structure of the IATS-D the specific factor for cardiovascular and respiratory processes seems to get lost. It may be that because cardiovascular and respiratory processes are more autonomic they are more distinctly noticeable when individuals think about how accurate they are in noticing these processes, but are less separable when individuals are reflecting on the attention they pay to these signals (i.e., they are part of a more general bodily awareness).

Both the IAS-D and IATS-D showed good internal consistency, comparable to those found in other general populations (Brand et al., 2023; Murphy et al., 2020; Tünte et al., 2024). However, in the factor structures some discrepancies were found. In the current study the CFA indicated a four-factor structure as the best model fit, which was different from previous findings. Along the same lines, the CFA model fit indicated a three-factor structure for the IATS-D. The three-factor structure was comparable to an English speaking sample (Gabriele et al., 2022), but differed from a German sample (Tünte et al., 2024). One explanation for these differing findings might be cultural differences. Even though people from the Netherlands, Germany, and the United Kingdom share similarities based on their Western European heritage, there are also several cultural differences that may influence study results. There might be differences in response styles (Harzing, 2006; van de Vijver & Leung, 2021), interpretation and translation of the questionnaires (Behr, 2017; Brislin, 1970), and cultural dimensions that impact underlying structures (Hofstede, 2001; Schwartz, 1999). For example, in terms of item interpretation, during the back-translation process, interviewees expressed difficulty in interpreting item 7 (tasting) as either being able to distinguish between different tastes (e.g., sweet, sour, bitter, etc.) or the ability to recognize when one food item tastes different from another food item. These subtle differences in interpretation of questions could have resulted in discrepancies in findings between the current and previous studies.

For the convergent, discriminant, and criterion validity, all our hypotheses were confirmed. Interoceptive accuracy showed significant negative associations with other measures of interoceptive accuracy, depressive symptoms, and alexithymia traits. This means that individuals who think they are accurate in perceiving their internal signals, are less likely to display depressive symptoms and alexithymia traits, and vice versa. A measure of interoceptive attention (i.e., BPQ-SF) did not determine interoceptive accuracy scores, indicating that self-reported accuracy of perceiving internal signals is not the same as paying attention to them. These findings mirror results from English and German versions of the IAS (Brand et al., 2023; Gabriele et al., 2022; Murphy et al., 2020; Tünte et al., 2024). Interoceptive attention was significantly and positively associated with other measures of interoceptive attention, interoceptive accuracy, depression, and alexithymia. The implication of these findings is that those who pay more attention to their bodily signals display more depressive symptoms and alexithymia traits, which is suggestive of maladaptive (i.e., insufficient or excessive attention to bodily signals, unfavorable for health-promoting behavior) forms of interoception (Trevisan et al., 2023). Again, these findings are in line with previous studies on the IATS in different cultural samples (Gabriele et al., 2022; Tünte et al., 2024). These differences between the IAS-D and the IATS-D signify that interoceptive accuracy and attention are distinctive interoceptive domains that interact with each other, and with depressive symptoms and alexithymia, in varying ways (Schoeller et al., 2025). Additionally, this shows that these questionnaires can distinguish between different interoceptive domains, and are valid instruments for measuring perceived interoception.

One of the strengths of this validation study was the large sample size, enhancing statistical power and robustness of the validation. The sample of 779 participants, with a diverse background in terms of age, region of the Netherlands where they reside, and socio-economic status ensured a fair representation of the Dutch population, contributing to the high external validity. Additionally, this is the first time the IAS and IATS were translated to Dutch and validated in a sample from the Netherlands. This allows for the new insights into the potential underlying structures of the IAS and IATS, as well as adding valuable knowledge on cultural differences in interoceptive processes. This study also has its limitations. First, this study had a cross-sectional design, and the questionnaires were not validated on test-retest reliability. This absence of a longitudinal aspect limits the conclusions that can be drawn about the stability of the IAS-D and IATS-D over longer periods of time. However, test-retest reliability is challenging because of learning effects: individuals may become more aware of their internal bodily signals every time they fill out a questionnaire, causing them to change their answers over time. Additionally, the self-report measures of interoception used in this study were not compared to objective interoceptive tasks, thus we are not able explain results in relation to these kind of tests. It is suggested that researchers should consider the complex relationship between objective and subjective interoceptive measures, in order to provide more accurate assessments (Schoeller et al., 2025). Second, we made use of a translated but not validated version of the ICQ. While this may have influenced the current findings, to be able to directly compare to previous studies using the same questionnaires the decision was made to use a back-translated version of the ICQ. The ICQ was translated using the same procedure as with the IAS and IATS, ensuring quality of translations and increasing confidence in the use of these measures. Finally, data collection in this study was performed using online surveys. For example, online testing may be limited because of selection bias (Bethlehem, 2010; Sue & Ritter, 2007) or participants rushing through surveys by providing random answers (Huang et al., 2012).

The model fit statistics of the CFA showed an imperfect fit for both the four-factor structure in the IAS-D (CFI = 0.816, TLI = 0.788, RMSEA = 0.084) and the three-factor structure in the IATS-D (CFI = 0.852, TLI = 0.831, RMSEA = 0.101). For the IAS-D this was comparable to other studies (Brand et al., 2023; Murphy et al., 2020, but for the IATS-D these values were lower in comparison to previous research (Tünte et al., 2024). Discrepancies in factor structures between the current and previous studies indicate the need for future research to further investigate the underlying structures and how the questionnaire items can be improved. Additionally, since test-retest reliability was not included in the current study, and no comparisons were made with objective interoceptive tests, the questionnaires could benefit from more longitudinal research in several interoceptive domains.

Nonetheless, the current study has shown that the IAS-D and IATS-D are valid measures of interoceptive accuracy and attention. The questionnaires can distinguish between these domains of interoception, and can possibly be used by practitioners to detect depressive symptoms and alexithymia. This is important, since it has been suggested that different interoceptive domains and modalities associate in varying ways with clinical symptoms (Schoeller et al., 2025). Finally, while some items may need refinement in order to improve interpretation, the overall questionnaires are easy to use, and are therefore relevant for use in practical settings.

In conclusion, the current study found that the IAS-D and IATS-D are reliable and valid instruments for the assessment of self-report interoceptive accuracy and attention in Dutch speaking populations. The results show that the questionnaires can distinguish between interoceptive domains, and are related to depression and alexithymia as expected from previous research. Future longitudinal research with these questionnaires in Dutch populations should focus on the association with objective interoception and test-retest reliability. These valid and reliable Dutch questionnaires offer accessible tools for assessing interoceptive accuracy and attention, supporting public health research in understanding the role of interoception in health behavior.

## Data Availability

All data, including the Dutch questionnaires, produced in the present study are available upon reasonable request to the authors

## 5. Other information

### Author contributions

JM, MEG, and JKJ conceptualized and designed the study. JM performed data collection. JM and JKJ conducted statistical analyses. JM, MEG, JdV, and JKJ contributed to the interpretation of the results. JM drafted the initial manuscript, with MEG, JdV, and JKJ providing critical revisions and refining the final manuscript. All authors reviewed and approved of the final version of the manuscript.

### Data availability

Questionnaires in Dutch, data, and analysis scripts will be made available by the corresponding author upon reasonable request.

### Declaration of generative AI and AI-assisted technologies in the writing process

During the preparation of this work the author(s) used ChatGPT in order to help with writing the code for the statistical analyses. After using this tool/service, the author(s) reviewed and edited the content as needed and take(s) full responsibility for the content of the published article.

## 7 Appendix

### Appendix A: Adjustments made after think-aloud interviews

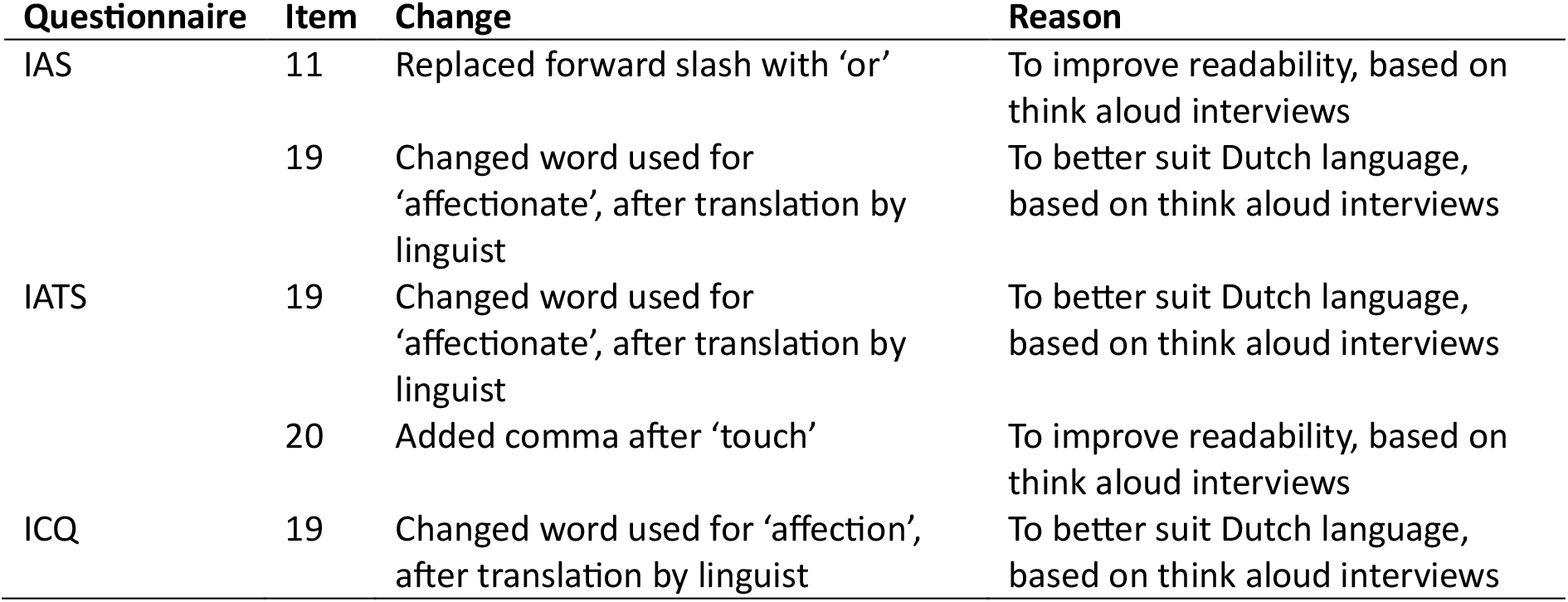

