## Supplemental File 1 for "Reliability and Validity of the Dutch Interoceptive Accuracy Scale and Interoceptive Attention Scale"

**Supplemental file 1: PCA factor loadings for the IAS-D and IATS-D.**

*Supplemental Table 1. Factor loadings for the Interoceptive Accuracy Scale Dutch (IAS-D) identified in the PCA.*

| Item | Factor 1 |
|---|---|
| Item 1: Heart | 0.139 |
| Item 2: Hunger | 0.232 |
| Item 3: Breathing | 0.178 |
| Item 4: Thirst | 0.198 |
| Item 5: Urination | 0.210 |
| Item 6: Defecation | 0.226 |
| Item 7: Tasting | 0.199 |
| Item 8: Vomiting | 0.222 |
| Item 9: Sneezing | 0.259 |
| Item 10: Coughing | 0.258 |
| Item 11: Temperature | 0.233 |
| Item 12: Sexual arousal | 0.213 |
| Item 13: Wind | 0.234 |
| Item 14: Burping | 0.248 |
| Item 15: Muscle soreness | 0.262 |
| Item 16: Bruising | 0.168 |
| Item 17: Pain | 0.244 |
| Item 18: Blood sugar | 0.109 |
| Item 19: Affective touch | 0.192 |
| Item 20: Tickling | 0.232 |
| Item 21: Itching | 0.254 |

*Supplemental Table 2. Factor loadings for the Interoceptive Attention Scale Dutch (IATS-D) identified in the PCA.*

| Item | Factor 1 |
|---|---|
| Item 1: Heart | 0.192 |
| Item 2: Hunger | 0.185 |
| Item 3: Breathing | 0.221 |
| Item 4: Thirst | 0.215 |
| Item 5: Urination | 0.209 |
| Item 6: Defecation | 0.247 |
| Item 7: Tasting | 0.089 |
| Item 8: Vomiting | 0.234 |
| Item 9: Sneezing | 0.251 |
| Item 10: Coughing | 0.251 |
| Item 11: Temperature | 0.222 |
| Item 12: Sexual arousal | 0.193 |
| Item 13: Wind | 0.238 |
| Item 14: Burping | 0.255 |
| Item 15: Muscle soreness | 0.238 |
| Item 16: Bruising | 0.221 |
| Item 17: Pain | 0.237 |
| Item 18: Blood sugar | 0.217 |
| Item 19: Affective touch | 0.138 |
| Item 20: Tickling | 0.214 |
| Item 21: Itching | 0.242 |
